# Age differences in clinical features and outcomes in patients with COVID-19, Jiangsu, China: a retrospective, multi-center cohort study

**DOI:** 10.1101/2020.06.01.20086025

**Authors:** Huanyuan Luo, Songqiao Liu, Yuancheng Wang, Penelope A. Phillips-Howard, Yi Yang, Shenghong Ju, Duolao Wang

## Abstract

**Objectives:** To determine the age-specific clinical presentations and incidence of adverse outcomes among patients with COVID-19 in Jiangsu, China.

**Design and setting:** This is a retrospective, multi-center cohort study performed at twenty-four hospitals in Jiangsu, China.

**Participants:** From January 10 to March 15, 2020, 625 patients with COVID-19 were involved.

**Results:** Of the 625 patients (median age, 46 years; 329 [52.6%] males), 37 (5.9%) were children (18 years or less), 261 (40%) young adults (19-44 years), 248 (39.7%) middle-aged adults (45-64 years), and 79 (12.6%) elderly (65 years or more). The incidence of hypertension, coronary heart disease, chronic obstructive pulmonary disease, and diabetes comorbidities increased with age (trend test, *P* <.0001, *P* = 0.0003, *P* <.0001, and *P* <.0001 respectively). Fever, cough, and shortness of breath occurred more commonly among older patients, especially the elderly, compared to children (Chi-square test, *P =* 0.0008, 0.0146, and 0.0282, respectively). The quadrant score and pulmonary opacity score increased with age (trend test, both *P* <.0001). Older patients had significantly more abnormal values in many laboratory parameters than younger patients. Elderly patients contributed the highest proportion of severe or critically-ill cases (33.0%, Chi-square test *P* < 0.001), intensive care unit (ICU) (35.4%, Chi-square test *P* < 0.001), and respiratory failure (31.6%, Chi-square test *P* = 0.0266), and longest hospital stay (21 days, ANOVA-test *P* < 0.001).

**Conclusions:** Elderly (≥65) patients with COVID-19 had the highest risk of severe or critical illness, intensive care use, respiratory failure, and the longest hospital stay, which may be due partly to that they had higher incidence of comorbidities and poor immune responses to COVID-19.

**Strengths and limitations of this study:** The cohort consists of almost all COVID-19 patients in Jiangsu province with a population over 80 million and its results should be representative of the patient population in the whole province and with a wide range of disease severity, therefore the results are subject to less selection bias.

The study includes imported and local cases and could study patients with different types of exposures.

The relative short follow-up time and a very small proportion of patients who remained in hospital after the 14-day follow-up period yield incomplete estimates for disease severity and clinical outcomes.

## Introduction

COVID-19 infection causes a wide spectrum of diseases which may lead to respiratory failure and death. Most of the available studies prior to the COVID-19 pandemic found that people of all ages are susceptible to SARS-CoV-2 infection but noted higher positive rates in real-time reverse transcriptase–polymerase chain reaction (RT-PCR) assays and hospitalization burden in older people.^1-3^ Similar to SARS, deaths and adverse clinical outcomes have been found to be more common in the elderly with known comorbidities for patients with COVID-19.^4-6^ Evidence suggests that asymptomatic carriers were more common among middle-aged people in close contact with infected family members.^7^ One study found that elderly patients with COVID-19 had some different clinical features from younger patients.^8^ Because age is a host factor that leads to a higher risk of severe COVID-19 and worse prognosis, it is important to better understand age-related susceptibility and pathology. However, published data on age differences in clinical features and clinical outcomes associated with COVID-19 are still scarce. This study aimed to investigate differences in clinical characteristics, disease severity, and clinical outcome burden in different age groups.

## Methods

### Study design and participants

This retrospective cohort study included all patients who met the patient’s inclusion and exclusion criteria, so no sample size calculations were conducted a priori. The inclusion criteria as of March 15, 2020, were all patients diagnosed with COVID-19 in Jiangsu Province according to the “Diagnosis and Treatment Protocol for Novel Coronavirus Pneumonia (Trial Version 7)” released by the National Health Commission & National Administration of Traditional Chinese Medicine of China;^9^ admitted to designated hospitals for COVID-19 treatment in Jiangsu. The exclusion criterion for patients was limited to those with no available medical records. The discharge standard imposed was that body temperature returns to normal for more than 3 days, symptoms resolve (if there were symptoms), and RT-PCR assays (throat swab samples, at least 1 day for sampling interval) show 2 consecutive negative results.

### Data collection and definition of variables

The epidemiological, clinical, laboratory, and radiologic parameters on admission; disease severity (asymptomatic, mild, moderate, severe, and critically ill); and clinical outcomes data were extracted from medical records. Data on disease severity were available at days 1, 2, 3, 4, 5, 6, 7 and 14 after admission, except for those who were discharged, and data on mortality and hospitalization status were available until March 15, 2020. Asymptomatic infection was defined as the absence of clinical symptoms but a positive nucleic acid test result. Mild disease was defined as having mild clinical symptoms and the absence of imaging manifestations of pneumonia in computer tomography (CT) scans. Moderate disease was defined as the presence of fever, respiratory tract symptoms or other symptoms and imaging manifestations of pneumonia. Severe disease was defined as the presence of at least one of the following items: respiratory distress, respiratory rate ≥ 30 beats / min; oxygen saturation in resting state (SpO_2_) ≤ 93%; or arterial blood oxygen partial pressure (PaO_2_) / fraction of inspired oxygen (FiO_2_) ≤ 300 mmHg (1 mmHg = 0.133kPa). Critically ill was defined as having respiratory failure requiring mechanical ventilation, shock or combined organ failure requiring intensive care unit (ICU) monitoring and treatment. We categorized the population into four age groups: children (18 years or less), young adults (19-44 years), middle-aged adults (45-64 years), and elderly (65 years or more).

All of the patients in Jiangsu had a high-resolution CT thorax examination which reflects lung lesions. CT images were assessed visually by two radiologists who had more than 5 years working experience in chest imaging. The radiologists were blinded to the patients’ information. Quadrant scores were the sum of the number of quadrants containing pulmonary opacities extending from the proximal to the distal end of the chest, with a score ranging between 0 and 4. For pulmonary opacity, bilateral lungs were scored manually and assigned an estimated percentage of pulmonary opacity relative to the whole lung, rounded to the nearest 5%.

### Statistical analysis

Continuous variables were reported as means ± standard deviation (SD) or median (interquartile range [IQR]) by groups and compared using ANOVA-test or Kruskal–Wallis test depending on their distributions. Categorical variables were summarized using frequency and percentage and compared using Chi-squared/Fisher exact test. To assess the linear trend effect of age on demographic and clinical variables and clinical outcomes, generalised linear models were employed with age (year) as the only predictor. Normal distribution and identity link function were used for continuous variables whereas binomial distribution and logit link function were used for binary variables. Analyses were performed using SAS 9.4 (SAS Institute), and a two-sided P < 0.05 was considered statistically significant.

### Patient and public involvement

Patients or the public were not involved in the design, or conduct, or reporting, or dissemination plans of our research.

## Results

Of the 721 suspected cases with possible COVID-19 during the study period, 631 patients were found to be RT-PCR positive for COVID-19. Only 625 patients were included in the study with complete data (Figure 1). The median age was 46 years (IQR, 32-57; range, 0.75-96 years), and 329 (52.6%) were men (Table 1). Thirty seven (5.9%) were children, 261 (40%) young adults of 19-44 years, 248 (39.7%) middle-aged of 45-64 years, and 79 (12.6%) elderly of 65 years or over.

**Figure 1:**
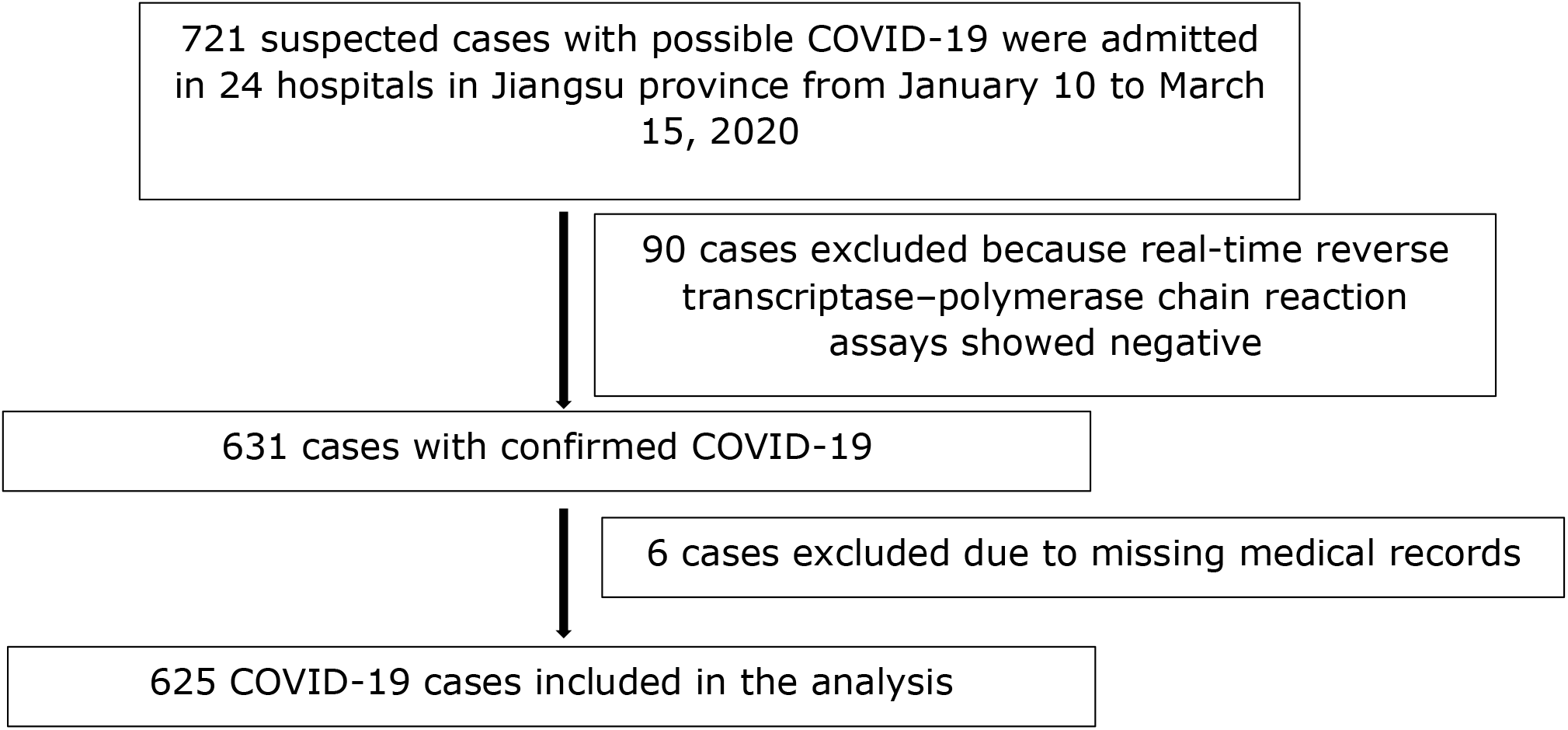
Study flow diagram

**Table 1:**
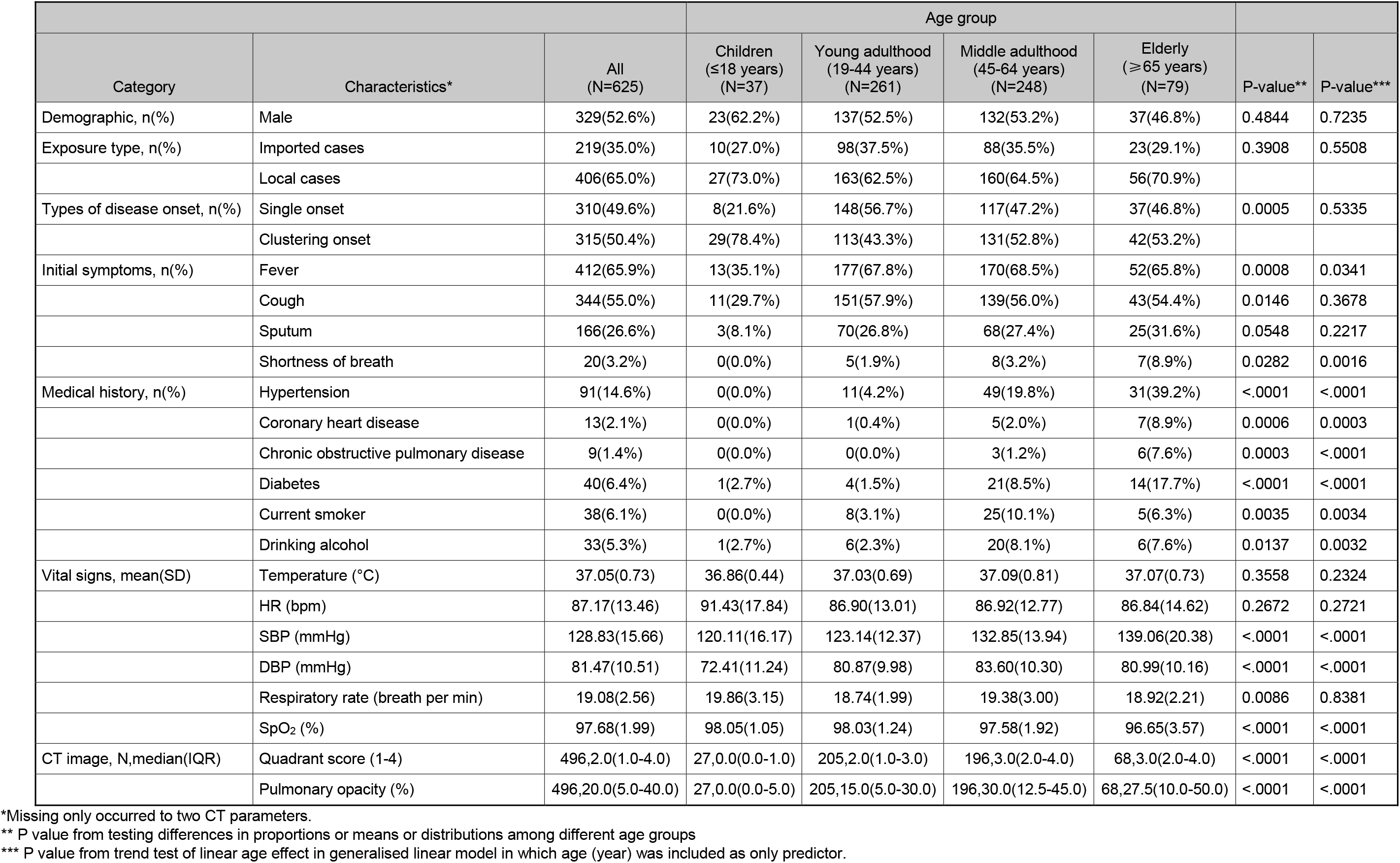
Demographic and clinical characteristics of patients with COVID-19 at admission by age group

| Category | Characteristics* | All<br>(N=625) | Age group |  |  |  | P-value** | P-value*** |
| --- | --- | --- | --- | --- | --- | --- | --- | --- |
|  |  |  | Children<br>(≤18 years)<br>(N=37) | Young adulthood<br>(19-44 years)<br>(N=261) | Middle adulthood<br>(45-64 years)<br>(N=248) | Elderly<br>(≥65 years)<br>(N=79) |  |  |
| Demographic, n(%) | Male | 329(52.6%) | 23(62.2%) | 137(52.5%) | 132(53.2%) | 37(46.8%) | 0.4844 | 0.7235 |
| Exposure type, n(%) | Imported cases | 219(35.0%) | 10(27.0%) | 98(37.5%) | 88(35.5%) | 23(29.1%) | 0.3908 | 0.5508 |
|  | Local cases | 406(65.0%) | 27(73.0%) | 163(62.5%) | 160(64.5%) | 56(70.9%) |  |  |
| Types of disease onset, n(%) | Single onset | 310(49.6%) | 8(21.6%) | 148(56.7%) | 117(47.2%) | 37(46.8%) | 0.0005 | 0.5335 |
|  | Clustering onset | 315(50.4%) | 29(78.4%) | 113(43.3%) | 131(52.8%) | 42(53.2%) |  |  |
| Initial symptoms, n(%) | Fever | 412(65.9%) | 13(35.1%) | 177(67.8%) | 170(68.5%) | 52(65.8%) | 0.0008 | 0.0341 |
|  | Cough | 344(55.0%) | 11(29.7%) | 151(57.9%) | 139(56.0%) | 43(54.4%) | 0.0146 | 0.3678 |
|  | Sputum | 166(26.6%) | 3(8.1%) | 70(26.8%) | 68(27.4%) | 25(31.6%) | 0.0548 | 0.2217 |
|  | Shortness of breath | 20(3.2%) | 0(0.0%) | 5(1.9%) | 8(3.2%) | 7(8.9%) | 0.0282 | 0.0016 |
| Medical history, n(%) | Hypertension | 91(14.6%) | 0(0.0%) | 11(4.2%) | 49(19.8%) | 31(39.2%) | <.0001 | <.0001 |
|  | Coronary heart disease | 13(2.1%) | 0(0.0%) | 1(0.4%) | 5(2.0%) | 7(8.9%) | 0.0006 | 0.0003 |
|  | Chronic obstructive pulmonary disease | 9(1.4%) | 0(0.0%) | 0(0.0%) | 3(1.2%) | 6(7.6%) | 0.0003 | <.0001 |
|  | Diabetes | 40(6.4%) | 1(2.7%) | 4(1.5%) | 21(8.5%) | 14(17.7%) | <.0001 | <.0001 |
|  | Current smoker | 38(6.1%) | 0(0.0%) | 8(3.1%) | 25(10.1%) | 5(6.3%) | 0.0035 | 0.0034 |
|  | Drinking alcohol | 33(5.3%) | 1(2.7%) | 6(2.3%) | 20(8.1%) | 6(7.6%) | 0.0137 | 0.0032 |
| Vital signs, mean(SD) | Temperature (°C) | 37.05(0.73) | 36.86(0.44) | 37.03(0.69) | 37.09(0.81) | 37.07(0.73) | 0.3558 | 0.2324 |
|  | HR (bpm) | 87.17(13.46) | 91.43(17.84) | 86.90(13.01) | 86.92(12.77) | 86.84(14.62) | 0.2672 | 0.2721 |
|  | SBP (mmHg) | 128.83(15.66) | 120.11(16.17) | 123.14(12.37) | 132.85(13.94) | 139.06(20.38) | <.0001 | <.0001 |
|  | DBP (mmHg) | 81.47(10.51) | 72.41(11.24) | 80.87(9.98) | 83.60(10.30) | 80.99(10.16) | <.0001 | <.0001 |
|  | Respiratory rate (breath per min) | 19.08(2.56) | 19.86(3.15) | 18.74(1.99) | 19.38(3.00) | 18.92(2.21) | 0.0086 | 0.8381 |
|  | SpO<sub>2</sub> (%) | 97.68(1.99) | 98.05(1.05) | 98.03(1.24) | 97.58(1.92) | 96.65(3.57) | <.0001 | <.0001 |
| CT image, N,median(IQR) | Quadrant score (1-4) | 496,2.0(1.0-4.0) | 27,0.0(0.0-1.0) | 205,2.0(1.0-3.0) | 196,3.0(2.0-4.0) | 68,3.0(2.0-4.0) | <.0001 | <.0001 |
|  | Pulmonary opacity (%) | 496,20.0(5.0-40.0) | 27,0.0(0.0-5.0) | 205,15.0(5.0-30.0) | 196,30.0(12.5-45.0) | 68,27.5(10.0-50.0) | <.0001 | <.0001 |

\*Missing only occurred to two CT parameters.
\*\* P value from testing differences in proportions or means or distributions among different age groups
\*\*\* P value from trend test of linear age effect in generalised linear model in which age (year) was included as only predictor.

There was no significant difference in the proportion of men and women in each age group (Table 1). A significantly higher proportion of young patients aged 18 years and below were cluster onset. The comorbidities incidence of hypertension, coronary heart disease, chronic obstructive pulmonary disease, and diabetes comorbidities increased with age (trend test, *P* <.0001, *P* = 0.0003, *P* <.0001, and *P* <.0001 respectively). Fever, cough, and shortness of breath occurred significantly more commonly among young adult, middle-aged, and elderly patients compared to children (67.8%, 68.5%, 65.8% vs 35.1%, *P* = 0.0008; 57.9%, 56%, 54.4%, vs 29.7%, *P* = 0.0146; 1.9%, 3.2%, 8.9%, vs 0%, *P* = 0.0282, respectively). The frequencies of these symptoms were similar among different age groups of adults except for shortness of breath which was dramatically more common (8.9%) in elderly adult patients of 65 years or over. Compared to children and younger adult patients, the incidence of smoking and drinking alcohol was significantly (twice the rate) higher among middle-aged and elderly patients. Three vital sign parameters on admission: SBP (mmHg), DBP (mmHg) and SpO_2_ (%), increased linearly with age (*P*<.0001 for three trend tests), with child patients having the lowest mean value. The quadrant score and pulmonary opacity score increased with age (trend tests, both *P* <.0001).

Significant differences were observed in some laboratory test results (Table 2). Older patients tended to have lower PaO2 (mmHg), PaCO2 (mmHg), and bicarbonate (mmol/L) in blood gas analysis. In blood tests they had lower white blood cell (WBC) (10^9^/L), lymphocyte (10^9^/L), hemoglobin (g/L), and platelets (10^9^/L). In organ function tests these patients had higher alanine aminotransferase (U/L) and creatinine (umol/L), and lower albumin (g/L). In coagulation function tests they had higher C-reactive protein (mg/L) in inflammatory factor tests; lower activated partial thromboplastin time (s), and higher fibrinogen (g/L) and d-dimer (mg/L).

**Table 2:** Laboratory parameters of patients with COVID-19 at admission by age group

|  |  |  | Age group, N, median(IQR) |  |  |  |  |  |
| --- | --- | --- | --- | --- | --- | --- | --- | --- |
| Category | Parameters | All<br>(N=625) | Children<br>(≤18 years)<br>(N=37) | Young adulthood<br>(19-44 years)<br>(N=261) | Middle adulthood<br>(45-64 years)<br>(N=248) | Elderly<br>(≥65 years)<br>(N=79) | P-value | P-value*** |
| Blood gas analysis | pH | 252,7.4(7.4-7.4) | 9,7.4(7.4-7.4) | 92,7.4(7.4-7.4) | 117,7.4(7.4-7.5) | 34,7.4(7.4-7.5) | 0.0213 | 0.0007 |
|  | PaO<sub>2</sub> (mmHg) | 252,94.0(78.0-110.0) | 9,94.0(86.7-113.0) | 92,102.0(87.0-124.5) | 117,89.0(76.2-106.0) | 34,81.7(74.0-101.0) | 0.0002 | 0.0009 |
|  | PaCO<sub>2</sub> (mmHg) | 252,39.0(36.0-42.0) | 9,45.0(42.0-49.0) | 92,39.5(37.2-42.0) | 117,39.0(35.6-42.0) | 34,37.2(34.6-42.0) | 0.0016 | 0.0087 |
|  | Lac (mmol/L) | 260,2.1(1.3-2.8) | 17,2.3(1.5-3.1) | 98,2.1(1.3-2.8) | 106,1.9(1.3-2.8) | 39,2.3(1.3-3.1) | 0.5056 | 0.5554 |
| Blood test | WBC Count (10<sup>9</sup>/L) | 513,4.9(3.9-6.2) | 33,6.0(4.8-7.2) | 213,4.9(3.9-6.1) | 205,4.8(3.8-6.1) | 62,4.6(3.9-6.4) | 0.0119 | 0.1275 |
|  | Neutrophil (10<sup>9</sup>/L) | 507,3.0(2.2-4.0) | 33,3.2(2.0-4.6) | 210,2.8(2.1-3.9) | 203,3.1(2.2-4.1) | 61,3.4(2.4-4.3) | 0.4466 | 0.2909 |
|  | Lymphocyte (10<sup>9</sup>/L) | 505,1.3(0.9-1.7) | 33,1.8(1.5-2.5) | 209,1.4(1.0-1.9) | 202,1.1(0.9-1.6) | 61,1.0(0.7-1.3) | <.0001 | <.0001 |
|  | Hemoglobin (g/L) | 510,137.0(124.0-150) | 33,138.0(126.0-148.0) | 212,139.5(123.0-154) | 204,136.5(125.0-148) | 61,129.0(121.0-140.0) | 0.0250 | 0.0050 |
|  | Platelet (10<sup>9</sup>/L) | 494,183.5(151.0-219) | 33,236.0(192.0-298.0) | 203,197.0(161.0-228) | 198,171.5(143.0-206) | 60,152.0(125.0-186.5) | <.0001 | <.0001 |
| Organ function | Alanine aminotransferase (U/L) | 425,25.0(16.5-38.0) | 33,19.0(16.0-30.0) | 169,25.0(16.0-39.3) | 170,26.8(19.0-42.0) | 53,22.0(15.0-33.0) | 0.0499 | 0.3126 |
|  | Albumin (g/L) | 480,41.4(38.0-45.1) | 33,45.0(40.1-47.9) | 194,42.1(38.0-46.6) | 193,41.5(38.0-44.4) | 60,38.9(35.4-43.1) | 0.0001 | <.0001 |
|  | Total bilirubin (umol/L) | 465,9.8(6.0-14.7) | 33,7.7(5.0-10.8) | 186,9.8(6.1-14.8) | 188,10.0(6.3-14.9) | 58,10.4(6.6-15.3) | 0.1141 | 0.3401 |
|  | Creatinine (umol/L) | 476,63.9(51.0-79.0) | 32,54.5(36.0-61.0) | 193,64.0(51.3-78.0) | 191,64.0(51.0-80.0) | 60,67.0(56.0-85.1) | 0.0003 | 0.0004 |
| Inflammatory factors | C-reactive protein (mg/L) | 474,10.0(2.7-22.6) | 30,5.1(1.2-10.0) | 193,9.6(2.0-16.2) | 189,10.0(4.4-26.3) | 62,16.8(5.0-51.1) | <.0001 | <.0001 |
|  | Procalcitonin (ng/mL) | 408,0.0(0.0-0.2) | 28,0.0(0.0-0.2) | 170,0.0(0.0-0.1) | 157,0.1(0.0-0.2) | 53,0.0(0.0-0.2) | 0.8557 | 0.0980 |
| Coagulation function test | Activated partial thromboplastin time (s) | 513,32.2(28.0-37.2) | 31,37.2(30.1-41.8) | 217,32.2(27.9-36.5) | 203,31.8(28.0-36.6) | 62,33.0(27.9-38.1) | 0.0448 | 0.2740 |
|  | Fibrinogen (g/L) | 496,3.5(2.7-4.2) | 30,2.6(2.2-2.9) | 209,3.3(2.6-4.1) | 198,3.8(3.0-4.4) | 59,3.7(2.6-4.8) | <.0001 | <.0001 |
|  | D-dimer (mg/L) | 475,0.2(0.1-0.4) | 30,0.2(0.1-0.3) | 191,0.2(0.1-0.3) | 195,0.2(0.1-0.4) | 59,0.4(0.2-0.7) | 0.0016 | 0.0049 |

\*\* P value from Kruskal-Wallis test of differences in distributions among different age groups
\*\*\* P value from trend test of linear age effect in generalised linear model in which age (year) was included as only predictor.

The proportion of patients who received supportive treatment and antiviral and antibiotic therapy increased significantly with patients’ age, except for the very rare procedure of continuous renal replacement therapy and extracorporeal membrane oxygenation treatment, and the very common use of interferon among different age groups (see supplementary Table S1).

The proportion of patients with severe or critical illness was 33% among elderly patients, compared with 13% among middle-aged patients, 2.3% among young adult patients, and 0% among children (*P* <.0001) (Table 3).

**Table 3:** Worst disease severity of patients with COVID-19 during hospital stay by age group

| Worst disease severity | Age group, n(%) |  |  |  |  | P-value |
| --- | --- | --- | --- | --- | --- | --- |
|  | All (N=625) | Children (≤18 years) (N=37) | Young adulthood (19-44 years) (N=261) | Middle adulthood (45-64 years) (N=248) | Elderly (≥65 years) (N=79) |  |
| Asymptomatic | 24(3.8%) | 8(21.6%) | 5(1.9%) | 7(2.8%) | 4(5.1%) | <.0001 |
| Mild | 34(5.4%) | 11(29.7%) | 18(6.9%) | 4(1.6%) | 1(1.3%) |  |
| Moderate | 503(80.5%) | 18(48.6%) | 232(88.9%) | 205(82.7%) | 48(60.8%) |  |
| Severe | 30(4.8%) | 0(0.0%) | 4(1.5%) | 16(6.5%) | 10(12.7%) |  |
| Critically ill | 34(5.4%) | 0(0.0%) | 2(0.8%) | 16(6.5%) | 16(20.3%) |  |

By the end of the study, none of the patients had died and all 625 patients had been discharged. The ICU rate (trend test, *P* < 0.001), respiratory failure rate (trend test, *P* < 0.001), and hospital stays (trend test, *P* < 0.001) increased with age (Table 4). The proportion of patients requiring ICU care (*P* < 0.001) and developing respiratory failure (*P* < 0.001) among elderly patients was 35.4% and 31.6% respectively, compared to 14.5% and 12.5% among middle-aged patients, 2.3% and 1.9% among young adult patients, and none among children, respectively. Elderly patients also had longer hospital stays (median [IQR], 21.0 [14.0-26.0] days) than all other age groups (15.0 [11.0-21.0] for children, 14.0 [11.0-19.0] for young adults, and 17.0 [13.0-22.0] for middle-aged adults, *P* <.0001).

**Table 4:** Clinical outcome of patients with COVID-19 by age group

| Clinical outcome | Age group, n(%) or median(IQR) |  |  |  |  |  |  |
| --- | --- | --- | --- | --- | --- | --- | --- |
|  | All (N=625) | Children (≤18 years) (N=37) | Young adulthood (19-44 years) (N=261) | Middle adulthood (45-64 years) (N=248) | Elderly (≥65 years) (N=79) | P-value* | P-value*** |
| ICU | 70(11.2%) | 0(0.0%) | 6(2.3%) | 36(14.5%) | 28(35.4%) | <.0001 | <.0001 |
| Shock | 2(0.3%) | 0(0.0%) | 0(0.0%) | 1(0.4%) | 1(1.3%) | 0.3581 | 0.0672 |
| Respiratory failure | 61(9.8%) | 0(0.0%) | 5(1.9%) | 31(12.5%) | 25(31.6%) | <.0001 | <.0001 |
| Renal failure | 2(0.3%) | 0(0.0%) | 0(0.0%) | 0(0.0%) | 2(2.5%) | 0.0031 | 0.0266 |
| Hospital stay (day) | 16.0(12.0-22.0) | 15.0(11.0-21.0) | 14.0(11.0-19.0) | 17.0(13.0-22.0) | 21.0(14.0-26.0) | <.0001 | <.0001 |

\*\* P value from testing differences in proportions or distributions among different age groups
\*\*\* P value from trend test of linear age effect in generalised linear model in which age (year) was included as only predictor.

## Discussion

We describe, to our knowledge, the largest cohort to date of 625 patients, to assess age differences in clinical features and clinical outcomes associated with COVID-19. We observed that most COVID-19 cases were among young adult patients of 19-44 years and middle-aged patients of 45–64 years, approximately 40% of each, with 5.9% children of 18 years or under, and 12.6% elderly patients of 65 years or over. This is consistent with a previous study from Korea reporting 6.3% of cases with COVID-19 were children under 19 years old as of March 20, which was considered to have tested the broadest and hence the most representative population.^10^

Our study showed that compared to adults, children were more likely to get infected through cluster gatherings. A previous study reported that COVID-19 in children was mainly caused by family transmission.^11^

Our study showed in Jiangsu, no patient had died and all patients had been discharged. Elderly patients were more than twice as likely to have a severe or critical illness compared with middle-aged patients, while young adult patients had a smaller proportion of severe symptoms, and child patients exhibited none. The ICU and respiratory failure rate, and hospital stay increased with age. Many case studies have shown that older patients are refractory (not yielding to treatment or not significantly improved after treatment) and likely to be at higher risk of more severe disease including acute respiratory distress syndrome (ARDS), respiratory failure, and death,^12-19^ while children are more likely to have mild or moderate type of COVID-19.^11,20^ This is similar to SARS characteristics that, compared to adults and adolescents, disease appears to be less severe in younger children.^6^

We found that initial symptoms including fever, cough, and shortness of breath occurred more frequently among adult patients than child patients and shortness of breath was dramatically more common in elderly adult patients of 65 years or over. This is consistent with the previous study showing the older group (≥60y) had a higher rate of shortness of breath than the younger group (<60y).^21^ We also found another study focused on age difference whose sample size (56 patients) was too small to draw a conclusion and only found 4 patients in total with symptoms of chest tightness or difficulty breathing.^8^ These symptoms may be the early signs of more severe illness and poorer outcomes in older patients. The frequency of these symptoms among adult patients in different age groups were similar except for shortness of breath, which was dramatically more common in elderly patients than younger age groups. This is different from the characteristics of influenza, where the initial clinical manifestations of frail elderly patients are usually subtle compared to young patients.^22,23^ In our study, the difference in vital sign of MAP on admission for different age groups were statistically significant (increasing with age) but may not be clinical significant.

Although early reports in Wuhan show that more men than women have severe COVID-19, later studies have reported a similar ratio of men and women in ICU.^24-26^ Although the inherent gender-immunity differences caused by chromosomes and hormones,^27^ as well as possible gender differences in smoking, drinking, and comorbidities, may have an impact on severity, the reason why earlier reports included more male patients, is probably that early patients were mainly males exposed in markets and congregation sites, and at higher risk for occupational infections.^26^ Our study shows that the proportion of men and women were similar for each age group, which may indicate that gender differences disappeared with the generalization of the study population.

Our study showed the age differences in clinical outcomes likely also resulted partially from the increased incidence of comorbidities with age including hypertension, coronary heart disease, chronic obstructive pulmonary disease, and diabetes, which may have increased the susceptibility of virus infection; such comorbidities are identified as a risk factor of more severe disease including respiratory failure and death in patients with COVID-19.^13,17-19,28^ Other explanations about why older people suffer poorer outcomes may be due to the higher prevalence of smoking and alcohol drinking in older patients with COVID-19 in Jiangsu. Our study showed seniors drank more than twice as often as young patients. History of smoking has been identified as a factor contributing to the progression of COVID-19 pneumonia.^29^

Our study demonstrated that the quadrant score and pulmonary opacity score increased with age, suggesting more severe abnormal imaging manifestation on admission among these older patients. This is consistent with the finding that the proportion of multiple lobe involvement in older patients was higher than in younger cases.^8^ Previous reports have also found some imaging differences by age groups; for example, primarily elderly patients were reported to have atypical imaging findings of consolidative opacities superimposed on ground-glass opacification,^30^ while paediatric patients showed more modest pulmonary involvement and less commonly reported consolidation complicated peripheral halo signs, compared with adults.^31-33^

Abnormal values in laboratory parameters in older patients may also be an early sign of, and a contribution to, severe illness and poor outcomes. This is consistent with a previous study showing the proportion of lymphocytes in the older cases was significantly lower and the C-reactive protein was higher than that in the younger patients.^8^ Studies showed albumin and C-reactive protein were associated with the progression of COVID-19 pneumonia,^29^ greater d-dimer on admission increases risk of in-hospital death,^14^ organ and coagulation dysfunction (eg, higher d-dimer) contributing to the development of ARDS and progression from ARDS to death.^16^

Our study indicated that lymphopenia (normal range: 0.3-3.0*10^9^/L) was shown in all age groups, but white blood cell and HYPERLINK “https://en.wikipedia.org/wiki/Lymphocyte” lymphocytes counts were lower in the older patients, indicating that live SARS-COV-2 virus stimulated poorer responses in the older patients. The mechanism in old patients with a severe COVID-19 illness may be that older patients have a diminished immune response to the novel SARS-COV-2 virus which is a defence mechanism against respiratory viruses and contributes to virus clearance. The thymic involution in older patients causes age-related reduction of T cell repertoire diversity and defects in CD4+ and CD8+ T cell function and hence significantly reduces immune function (known as immunosenescence).^34,35^ Immunosenescence makes many viral infections worse in older patients.^36^ But further research is necessary to investigate whether age differences in disease severity and outcomes of COVID-19 result from aging of the immune system and reduced responsiveness. This is because some respiratory viruses could escape antiviral mechanisms and immune responses.^37^ For example, a study of H1N1pdm (respiratory viruses) on ferrets found no significant difference in viral clearance between young and adult subjects.^38^ However, studies found that the pulmonary pathology improved earlier in young ferrets, regulatory interleukin-10 (which is mainly produced by monocytes and lymphocytes) and interferon responses were more robust in young ferrets.^38-40^ Also H1N1pdm infection triggered formation of lung structures that resembled inducible bronchus-associated lymphoid tissues (iBALTs) in young ferrets which contributes to pulmonary immune responses and were not seen in the adult ferrets with severe disease.^38-40^ Some other studies demonstrated aged ferrets infected with influenza viruses had reduced antibody production and delayed peripheral blood T-cell responses compared to adult comterparts.^34,35,41^ Another study reported that except for immunosenescence in aged people, age-related increases in levels of phospholipase could also result in a delay of immune response and poor outcomes after SARS-CoV infection.^36^ Overall, research focused on innate immune-related mechanisms and viral clearance in patients with COVID-19 of different age groups may help determine the underlying mechanisms of disease severity.

Some other reasonable mechanisms of mild presentation in children include qualitatively different response to SARS-CoV2, or different expression of angiotensin-converting enzyme (ACE) 2 receptors required for SARS-CoV2 infection, or different virus-to-virus interaction and competition from other viruses limiting SARS-CoV2 growth.^10^

A larger proportion of older patients in this study received supportive treatment and antiviral and antibiotic therapies which was due in part to the increasing proportion of severely or critically illness in older adults. A previous study similarly showed that treatment was statistically different by age group.^8^

We believe that the findings of this study are generalizable to populations in similar settings (e.g. outside the initial pandemic center) for three reasons: (1) we included all patients who met the inclusion and exclusion criteria during the study period; (2) the study population consisted of cases confirmed by laboratory tests, by screening suspected cases especially those who had a contact of pandemic center, or a contact of people who had been to pandemic center or who had confirmed diagnoses of COVID-19; and (3) after the end of study, no new cases was confirmed with COVID-19 because of the effective and timely public health interventions including isolation of suspected and confirmed cases and lockdown of the whole country.

Our study has several limitations. First, the relative short follow-up time and a very small proportion of patients who remained in hospital after the 14-day follow-up period yield incomplete estimates for disease severity and clinical outcomes, making it difficult to fully assess age differences in the burden associated with COVID-19. However, this impact is minor and may not strongly affect the study results because we included analyses of outcomes at the end of study, and only a small number of patients were still in hospital at the end of the study. Second, we were unable to perform multiple regression analysis to control for possible bias in the observed age impact in clinical features and outcomes. As a result, the observed age differences may still be subject to possible confounding factors.

## Conclusions

Older patients in this setting in China tended to have relatively severe clinical infections and poor clinical outcomes associated with COVID-19 compared to younger patients. Elderly patients aged 65 and over were at a much higher risk of developing severe or critical illness than other age groups. The ICU and respiratory failure rate, and hospital stay increased with age. Older patients have worse clinical outcomes, in part due to comorbidities in older people, and higher rates of smoking and drinking habits, and immune, organ, and coagulation dysfunction on admission. In studying the pathogenesis and developing management strategies of COVID-19, the age factor is confirmed as a critical factor in severity of infection.

## Data Availability

All data are freely available within the appendices. No additional data available.

## Contributors

DW, SJ and YY conceived and designed the study. HL, SL, and YW contributed to the literature search. SL, YW, SJ, YY, and DW contributed to data collection, quality checks and data management. DW, HL, SJ, SL, PPH, YW and YY contributed to data analysis and results presentation. DW, HL, SL, SJ, PPH, and YY were responsible for results interpretation. HL, DW, PPH, SL, YW, SJ, SL and YY contributed in the drafting and review of the manuscript.

## Ethical approval

The study was approved by the Ethics Committee of Zhongda Hospital Affiliated to Southeast University (2020ZDSYLL013-P01 and 2020ZDSYLL019-P01).

## Patient consent

Informed consent was waived due to the emergent pandemic.

## Data sharing

All data are freely available within the appendices. No additional data available.

## Reference

1. Wang S, Guo L, Chen L, et al. A case report of neonatal COVID-19 infection in China. Clin Infect Dis. 2020. pii: ciaa225. doi: 10.1093/cid/ciaa225. [Epub ahead of print].

2. Sun D, Li H, Lu XX, et al. Clinical features of severe pediatric patients with coronavirus disease 2019 in Wuhan: a single center’s observational study. World J Pediatr. 2020. doi: 10.1007/s12519-020-00354-4. [Epub ahead of print].

3. Liu R, Han H, Liu F, et al. Positive rate of RT-PCR detection of SARS-CoV-2 infection in 4880 cases from one hospital in Wuhan, China, from Jan to Feb 2020. Clin Chim Acta. 2020;505:172–175.

4. Porcheddu R, Serra C, Kelvin D, Kelvin N, Rubino S. Similarity in Case Fatality Rates (CFR) of COVID-19/SARS-COV-2 in Italy and China. J Infect Dev Ctries. 2020;14(2):125–128.

5. Wong RS, Wu A, To KF, et al. Haematological manifestations in patients with severe acute respiratory syndrome: retrospective analysis. Bmj. 2003;326(7403):1358–1362.

6. Hon KL, Leung CW, Cheng WT, et al. Clinical presentations and outcome of severe acute respiratory syndrome in children. Lancet. 2003;361(9370):1701–1703.

7. Wang Y, Liu Y, Liu L, Wang X, Luo N, Ling L. Clinical outcome of 55 asymptomatic cases at the time of hospital admission infected with SARS-Coronavirus-2 in Shenzhen, China. J Infect Dis. 2020. pii: jiaa119. doi: 10.1093/infdis/jiaa119. [Epub ahead of print].

8. Liu K, Chen Y, Lin R, Han K. Clinical feature of COVID-19 in elderly patients: a comparison with young and middle-aged patients. J Infect. 2020. pii: S0163–4453(20)30116-X. doi: 10.1016/j.jinf.2020.03.005. [Epub ahead of print].

9. National Health Commission & National Administration of Traditional Chinese Medicine. Diagnosis and Treatment Protocol for Novel Coronavirus Pneumonia (Trial Version 7). Chin Med J (Engl). 2020;133. doi: 10.1097/CM9.0000000000000819.

10. Brodin P. Why is COVID-19 so mild in children? Acta Paediatr. 2020. doi: 10.1111/apa.15271. [Epub ahead of print].

11. Su L, Ma X, Yu H, et al. The different clinical characteristics of corona virus disease cases between children and their families in China - the character of children with COVID-19. Emerg Microbes Infect. 2020;9(1):707–713.

12. Mo P, Xing Y, Xiao Y, et al. Clinical characteristics of refractory COVID-19 pneumonia in Wuhan, China. Clin Infect Dis. 2020. pii: ciaa270. doi: 10.1093/cid/ciaa270. [Epub ahead of print].

13. Yang X, Yu Y, Xu J, et al. Clinical course and outcomes of critically ill patients with SARS-CoV-2 pneumonia in Wuhan, China: a single-centered, retrospective, observational study. Lancet Respir Med. 2020. pii: S2213–2600(20)30079-5. doi: 10.1016/S2213-2600(20)30079-5. [Epub ahead of print].

14. Zhou F, Yu T, Du R, et al. Clinical course and risk factors for mortality of adult inpatients with COVID-19 in Wuhan, China: a retrospective cohort study. Lancet. 2020. pii: S0140–6736(20)30566-3. doi: 10.1016/S0140-6736(20)30566-3. [Epub ahead of print].

15. Shim E, Tariq A, Choi W, Lee Y, Chowell G. Transmission potential and severity of COVID-19 in South Korea. Int J Infect Dis. 2020. pii: S1201–9712(20)30150-8. doi: 10.1016/j.ijid.2020.03.031. [Epub ahead of print].

16. Wu C, Chen X, Cai Y, et al. Risk Factors Associated With Acute Respiratory Distress Syndrome and Death in Patients With Coronavirus Disease 2019 Pneumonia in Wuhan, China. JAMA Intern Med. 2020. doi: 10.1001/jamainternmed.2020.0994. [Epub ahead of print].

17. Liu K, Fang YY, Deng Y, et al. Clinical characteristics of novel coronavirus cases in tertiary hospitals in Hubei Province. Chin Med J (Engl). 2020. doi: 10.1097/CM9.0000000000000744. [Epub ahead of print].

18. Lai CC, Liu YH, Wang CY, et al. Asymptomatic carrier state, acute respiratory disease, and pneumonia due to severe acute respiratory syndrome coronavirus 2 (SARS-CoV-2): Facts and myths. J Microbiol Immunol Infect. 2020. pii: S1684–1182(20)30040-2. doi: 10.1016/j.jmii.2020.02.012. [Epub ahead of print].

19. Guo YR, Cao QD, Hong ZS, et al. The origin, transmission and clinical therapies on coronavirus disease 2019 (COVID-19) outbreak - an update on the status. Mil Med Res. 2020;7(1):11.

20. Qiu H, Wu J, Hong L, Luo Y, Song Q, Chen D. Clinical and epidemiological features of 36 children with coronavirus disease 2019 (COVID-19) in Zhejiang, China: an observational cohort study. Lancet Infect Dis. 2020. pii: S1473–3099(20)30198-5. doi: 10.1016/S1473-3099(20)30198-5. [Epub ahead of print].

21. Lian J, Jin X, Hao S, et al. Analysis of Epidemiological and Clinical features in older patients with Corona Virus Disease 2019 (COVID-19) out of Wuhan. Clin Infect Dis. 2020. pii: ciaa242. doi: 10.1093/cid/ciaa242. [Epub ahead of print].

22. Pop-Vicas A, Gravenstein S. Influenza in the elderly: a mini-review. Gerontology. 2011;57(5):397–404.

23. Wong PL, Sii HL, P’Ng CK, et al. The effects of age on clinical characteristics, hospitalization and mortality of patients with influenza-related illness at a tertiary care centre in Malaysia. Influenza Other Respir Viruses. 2020. doi: 10.1111/irv.12691. [Epub ahead of print].

24. Chen N, Zhou M, Dong X, et al. Epidemiological and clinical characteristics of 99 cases of 2019 novel coronavirus pneumonia in Wuhan, China: a descriptive study. Lancet. 2020;395(10223):507–513.

25. Huang C, Wang Y, Li X, et al. Clinical features of patients infected with 2019 novel coronavirus in Wuhan, China. Lancet. 2020;395(10223):497–506.

26. Wang D, Hu B, Hu C, et al. Clinical Characteristics of 138 Hospitalized Patients With 2019 Novel Coronavirus-Infected Pneumonia in Wuhan, China. JAMA. 2020. doi: 10.1001/jama.2020.1585. [Epub ahead of print].

27. 308–321 JSSdiiiCRAI. Sexual dimorphism in innate immunity. Clin Rev Allergy Immunol. 2019;56:308-321.

28. Leung C. Clinical features of deaths in the novel coronavirus epidemic in China. Rev Med Virol. 2020:e2103.

29. Liu W, Tao ZW, Lei W, et al. Analysis of factors associated with disease outcomes in hospitalized patients with 2019 novel coronavirus disease. Chin Med J (Engl). 2020. doi: 10.1097/CM9.0000000000000775. [Epub ahead of print].

30. Salehi S, Abedi A, Balakrishnan S, Gholamrezanezhad A. Coronavirus Disease 2019 (COVID-19): A Systematic Review of Imaging Findings in 919 Patients. AJR Am J Roentgenol. 2020: 1–7.

31. Xia W, Shao J, Guo Y, Peng X, Li Z, Hu D. Clinical and CT features in pediatric patients with COVID-19 infection: Different points from adults. Pediatr Pulmonol. 2020. doi: 10.1002/ppul.24718. [Epub ahead of print].

32. Liu H, Liu F, Li J, Zhang T, Wang D, Lan W. Clinical and CT imaging features of the COVID-19 pneumonia: Focus on pregnant women and children. J Infect. 2020. pii: S0163–4453(20)30118-3. doi: 10.1016/j.jinf.2020.03.007. [Epub ahead of print].

33. Li W, Cui H, Li K, Fang Y, Li S. Chest computed tomography in children with COVID-19 respiratory infection. Pediatr Radiol. 2020. doi: 10.1007/s00247-020-04656-7. [Epub ahead of print].

34. Maue AC, Yager EJ, Swain SL, Woodland DL, Blackman MA, Haynes L. T-cell immunosenescence: lessons learned from mouse models of aging. Trends Immunol. 2009;30(7):301–305.

35. Yager EJ, Ahmed M, Lanzer K, Randall TD, Woodland DL, Blackman MA. Age-associated decline in T cell repertoire diversity leads to holes in the repertoire and impaired immunity to influenza virus. J Exp Med. 2008;205(3):711–723.

36. Vijay R, Hua X, Meyerholz DK, et al. Critical role of phospholipase A2 group IID in age-related susceptibility to severe acute respiratory syndrome-CoV infection. J Exp Med. 2015;212(11):1851–1868.

37. Vareille M, Kieninger E, Edwards MR, Regamey N. The airway epithelium: soldier in the fight against respiratory viruses. Clin Microbiol Rev. 2011;24(1):210–229.

38. Huang SS, Banner D, Degousee N, et al. Differential pathological and immune responses in newly weaned ferrets are associated with a mild clinical outcome of pandemic 2009 H1N1 infection. J Virol. 2012;86(24):13187–13201.

39. Hwang JY, Randall TD, Silva-Sanchez A. Inducible Bronchus-Associated Lymphoid Tissue: Taming Inflammation in the Lung. Front Immunol. 2016;7:258.

40. Randall TD. Bronchus-associated lymphoid tissue (BALT) structure and function. Adv Immunol. 2010;107:187–241.

41. Paquette SG, Huang SSH, Banner D, et al. Impaired heterologous immunity in aged ferrets during sequential influenza A H1N1 infection. Virology. 2014;464–465:177-183.

